# Clinical Validation of the EMOCARE-Derived Depressive Symptom Severity Score using Established Clinician- and Self-reported Scales: Preliminary Evidence Across 3 Prospective Studies

**DOI:** 10.64898/2026.03.08.26347894

**Authors:** Antony Perzo, Tanel Petelot, Renaud Seguier, Igor Magaraggia

## Abstract

In a pooled analysis of 3 prospective studies in adults with mood disorders, EMOCARE depressive symptom severity scores derived from passive multimodal remote monitoring showed moderate-to-strong convergent validity and sensitivity to change versus established clinician- and self-rated symptom scales, including the Montgomery-Åsberg Depression Rating Scale (MADRS) and Patient Health Questionnaire-9 (PHQ-9).

## Introduction

Passive remote-monitoring digital health technologies (DHTs) may complement episodic symptom scales by enabling higher-frequency, lower-burden assessment of symptom dynamics [1]. For broad clinical and regulatory uptake, passively derived measures should demonstrate fit-for-purpose validity against established clinical anchors [2,3]. EMOCARE is a passive multimodal DHT that derives depressive symptoms severity scores from smartphone-based sensing (e.g., facial/voice-derived features, activity proxies, and digital behavior) and presents longitudinal trajectories for clinical review [4]. We report pooled interim evidence of convergent validity and sensitivity to change of EMOCARE versus golden standard symptom rating scales across 3 prospective clinical studies.

## Methods

### Study design and data sources

We pooled observations from 3 prospective observational studies in Major Depressive Disorder (EMC1: ID-RCB 2024-A01487-40; EMC2FR: ID-RCB 2024-A02120-47), and Bipolar Disorder (EMC2-BD: ID-RCB 2025-A00100-49). Each protocol included repeated symptom assessments aligned with concurrent passive monitoring. Participants provided written informed consent before enrollment, per protocol. Where applicable, this manuscript follows relevant STROBE and DECIDE-AI reporting guidelines (see Multimedia Appendix for completed checklists).

### EMOCARE score derivation

EMOCARE software (EMOBOT, France) passively acquires multimodal inputs during routine device use (e.g., opportunistic front-camera snapshots, short audio snippets when voice activity is detected, actigraphy/motion signals, and digital behavior such as screen/unlock and session metrics) and transforms these signals into quantitative behavioral features. Derived features are synchronized in encrypted form and aggregated server-side to compute a depressive symptoms severity score (i.e., EMOCARE score) on a 0–100 scale, updated daily in a rolling fashion as additional data accrue. EMOCARE score availability depends on “valid-day” criteria and modality coverage rules; for initial score generation, the device description specifies a requirement of ≥7 valid days within a 14-day window, where a day is considered valid when ≥800 EMOCARE data points/day are collected. Importantly, the AI model underlying the EMOCARE software is ‘frozen’ since 2023. Consequently, the EMOCARE score is identically derived across all three studies (same preprocessing pipelines, and quality filtering procedures).

### Clinical anchors

Reference anchors for assessing depressive symptomatology included both clinician-reported (i.e., MADRS, HAM-D17), and patient-reported scales (i.e., PHQ-9, BDI-II, GAD-7), with well-established validity and regulatory recognition. Depending on the study, these scales were administered at baselines and various follow-up visits, either remotely or in-clinic. Pooling of data was performed exclusively with reference scales administered in a consistent manner across studies.

### Statistical analysis

Within-person concordance across repeated observations was quantified via repeated-measures correlation (rmCorr), requiring ≥3 paired observations per participant. Concurrent validity was evaluated using Spearman rank correlation (ρ). Sensitivity to change used Spearman correlation between consecutive-visit changes (ΔEMOCARE score vs Δanchor) among consecutive valid visits. To reflect longitudinal clustering (multiple measurements/person), 95% CIs were estimated using participant-level bootstrap resampling (bias-corrected and accelerated), retaining all rows for each resampled participant, and P values were generated via cluster-wise permutation testing (50,000 shuffles) that preserved within-participant structure. Statistical analyses were performed using X version A.

## Results

Across pooled analyzable observations, EMOCARE scores showed moderate-to-strong concurrent associations with symptom scales (ρ range 0.613–0.833). Within-person concordance was strong for MADRS (rmCorr r=0.895) and moderate-to-strong for PHQ-9 and GAD-7. EMOCARE also demonstrated sensitivity to change, with strong correlation to PHQ-9 change (ρ=0.834) and moderate-to-strong correlation to GAD-7 change (ρ=0.655; Table 1).

**Table 1.**
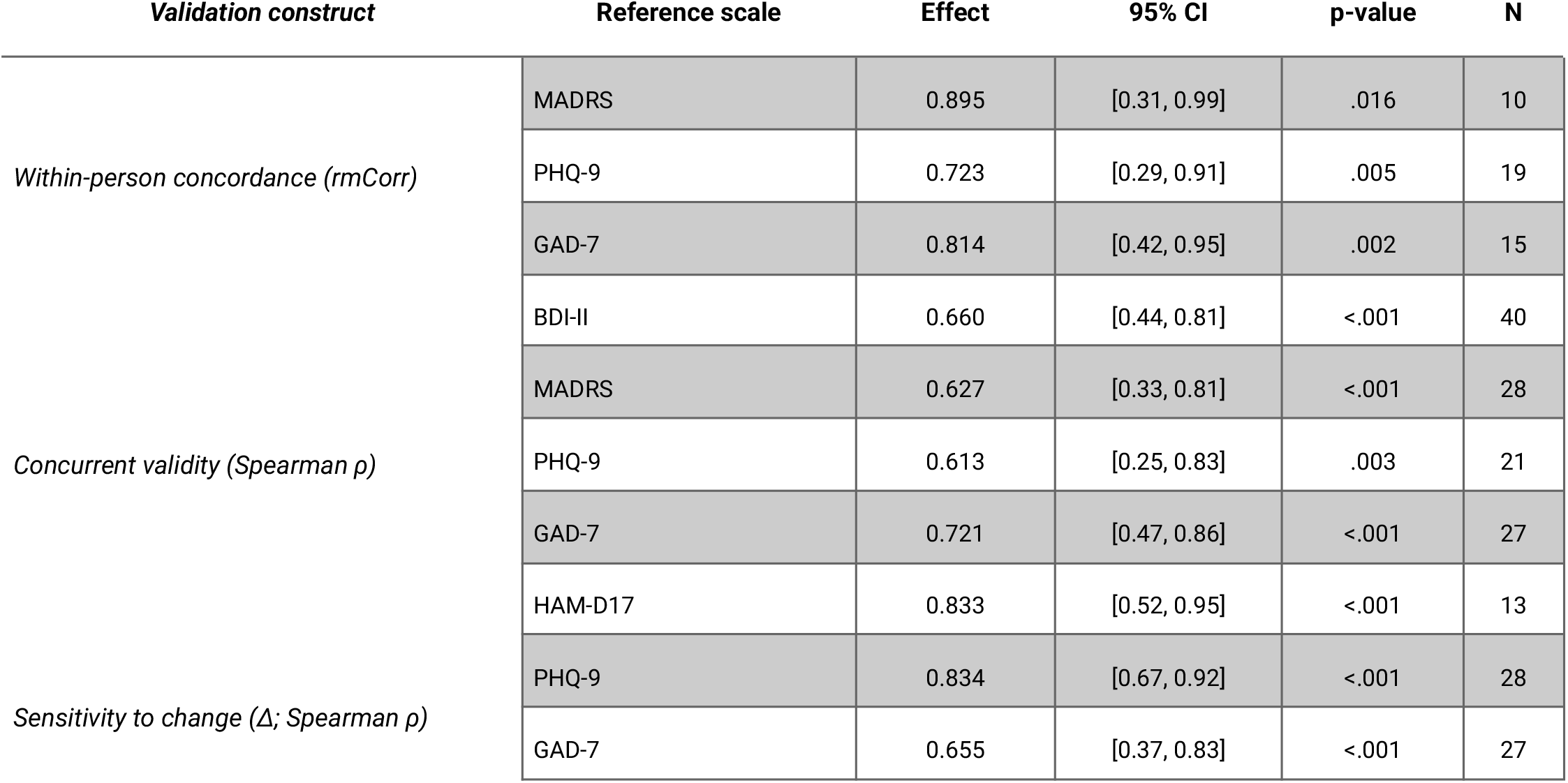
Pooled convergent validity and sensitivity-to-change results for EMOCARE versus reference scales. For Spearman and rmCorr analyses, N denotes participants with paired observations and consecutive-visit pairs, respectively.

## Discussion

In pooled interim data from 3 prospective studies in adults with mood disorders, EMOCARE demonstrated concurrent/convergent validity against multiple clinician- and self-rated symptom scales and was responsive to symptom change over time. As such, this work addresses one key evidentiary component for adoption of DHT-derived endpoints: interpretability relative to accepted anchors, rather than prediction of discrete clinical events or demonstration of clinical utility [1,3].

A notable pattern in these pooled results is that EMOCARE’s associations are not uniform across clinician-rated versus patient-reported anchors, which is unsurprising—and scientifically informative—given that these instruments are not interchangeable measures of a single latent construct. Clinician-rated scales (e.g., MADRS, HAM-D17) explicitly integrate observable and interview-elicited phenomena (psychomotor change, affective expression, speech latency, sleep/appetite as described in context), many of which plausibly overlap with EMOCARE’s passively derived behavioral proxies (e.g., activity, vocal/visual features, device interaction) [5,6]. This can inflate convergent validity by genuine construct overlap, but it also raises a critical point: clinician ratings may partially reflect the same outward cues that EMOCARE quantifies, reducing independence of the comparator. In contrast, patient-reported scales (e.g., PHQ-9, BDI-II) place heavier weight on internal experiences (rumination, guilt, hopelessness, worry, subjective distress) and are susceptible to recall context, insight, and response style [7–9]; these dimensions may be only weakly expressed in passive behavioral channels, yielding lower correlations that reflect true construct non-overlap rather than “poor performance.” Additional contributors include temporal misalignment (heterogeneous recall windows vs a 14-day EMOCARE aggregation that smooths short-lived fluctuations), differential measurement error (unsupervised self-report vs structured interviews), and pooling-induced range restriction and heterogeneity (different sites, schedules, and diagnoses). Together, cross-scale differences should be interpreted as mapping the boundary conditions of what EMOCARE captures.

Several limitations inherent to this study warrant consideration. First, pooling across protocols introduces heterogeneity in diagnosis (MDD vs bipolar disorder), follow-up cadence, and anchor availability. Second, the analyzable dataset is defined by an a priori passive-data density requirement (≥7 valid days/14 days; ≥800 data points/day), which strengthens measurement integrity but may induce selection effects if adherence, device usage patterns, or permission settings differ systematically across participants. Third, within-person rmCorr analyses required ≥3 paired observations per participant, resulting in smaller Ns and wider uncertainty for those estimates. Fourth, some anchors (e.g., PHQ-9) have 2-week recall windows, which may partially overlap the 14-day EMOCARE window by design; however, other instruments and visit schedules may yield imperfect temporal alignment, potentially attenuating correlations. Fifth, multiple correlations were examined across different scales; this interim report emphasizes effect sizes with uncertainty estimates rather than formal multiplicity control.

Overall, these pooled interim findings provide early evidence that passive multimodal monitoring with EMOCARE yields severity estimates that are concordant with established symptom scales and responsive to symptom dynamics. Future work should align and document validity/data-density definitions across device documentation and analysis, and evaluate clinically meaningful change thresholds and broader generalizability under varied real-world operating conditions.

## Data Availability

The datasets analyzed during this study are not publicly available due to privacy and confidentiality constraints but are available from the corresponding author on reasonable request.

## Acknowledgements

The authors gratefully acknowledge the non-financial contributions of colleagues and institutions whose support did not meet the ICMJE criteria for authorship, including…

## Funding Statement

This work was sponsored by Emobot SAS.

## Conflicts of Interest

Emobot is the manufacturer of EMOCARE and the sponsor of the aforementioned studies.

## Author Contributions

Antony Perzo: Conceptualization; Methodology; Formal analysis; Investigation; Writing – original draft; Writing – review & editing; Visualization; Supervision; Project administration Tanel Petelot: Conceptualization; Methodology; Writing – review & editing; Supervision; Project administration Renaud Séguier: Writing – review & editing Igor Magaraggia: Writing – original draft; Writing – review & editing; Supervision

## Abbreviations

BDI-II: Beck Depression Inventory-II
DHT: digital health technology
GAD-7: Generalized Anxiety Disorder-7
HAM-D17: Hamilton Depression Rating Scale-17
MADRS: Montgomery-Åsberg Depression Rating Scale
MDD: major depressive disorder
PHQ-9: Patient Health Questionnaire-9
rmCorr: repeated-measures correlation

## Notes

### Author Declarations

Comite de Protection des Personnes Sud-Est I of the Centre Hospitalier Universitaire de Saint-Etienne gave ethical approval for this work.

